# Integrative Systems Immunology Analysis Reveals Elevated Anti-AGTR1 Levels with Accumulating COVID-19 Symptoms

**DOI:** 10.1101/2024.04.05.24305287

**Authors:** Dennyson Leandro M Fonseca, Maj Jäpel, Igor Salerno Filgueiras, Gabriela Crispim Baiochi, Yuri Ostrinski, Gilad Halpert, Yael Bublil Lavi, Elroy Vojdani, Juan Carlo Santos e Silva, Júlia Nakanishi Usuda, Paula P. Freire, Adriel Leal Nóbile, Anny Silva Adri, Pedro Barcelos Marçal, Yohan Lucas Gonçalves Corrêa, Fernando Yuri Nery do Vale, Letícia Oliveira Lopes, Solveig Lea Schmidt, Xiaoqing Wang, Carl Vahldieck, Benedikt Fels, Lena F. Schimke, Mario Hiroyuki Hirata, Gustavo Cabral- Miranda, Taj Ali AKhan, Rusan Catar, Guido Moll, Thayna Silva-Sousa, Yen-Rei A Yu, Rodrigo JS Dalmolin, Howard Amital, Aristo Vojdani, Helder Nakaya, Hans D. Ochs, Jonathan I. Silverberg, Jason Zimmerman, Israel Zyskind, Avi Z Rosenberg, Kai Schulze-Forster, Harald Heidecke, Alexander Hackel, Kristina Kusche-Vihrog, Yehuda Shoenfeld, Gabriela Riemekasten, Reza Akbarzadeh, Alexandre H.C Marques, Otavio Cabral-Marques

## Abstract

The coronavirus disease 2019 (COVID-19) displays a broad spectrum of symptoms, with the underlying reasons for this variability still not fully elucidated. Our study investigates the potential association between specific autoantibodies (AABs), notably those that targeting G protein-coupled receptors (GPCRs) and renin-angiotensin system (RAS) related molecules, and the diverse clinical manifestations of COVID-19, commonly observed in patients with autoimmune conditions, including rheumatic diseases, such as systemic sclerosis. In a cross-sectional analysis, we explored the relationship between AAB levels and the presence of key COVID-19 symptoms. Hierarchical clustering analysis revealed a robust correlation between certain AABs and symptoms such as fever, muscle ache, anosmia, and dysgeusia, which emerged as significant predictors of disease severity. Specifically, AABs against CHRM5 and CXCR3 were strongly linked to fever, while AABs against CHRM5 and BDKRB1 correlated with muscle ache. Anosmia was predominantly associated with AABs against F2R and AGTR1, while dysgeusia was linked to AABs against BDKRB1 and AGTR1. Furthermore, we observed a rise in AAB levels with the accumulation of these symptoms, with the highest levels detected in patients presenting all four predictors. Multinomial regression analysis identified AABs targeting AGTR1 as a key predictor for one or more of these core symptoms. Additionally, our study indicated that anti-AGTR1 antibodies triggered a concentration-dependent degradation of eGC, which could be mitigated by the AGTR1 antagonist Losartan. This suggests a potential mechanistic connection between eGC degradation, the observed COVID-19 symptoms, and rheumatic diseases. In conclusion, our research underscores a substantial correlation between AABs, particularly those against GPCRs and RAS-related molecules, and the severity of COVID-19 symptoms. These findings open avenues for potential therapeutic interventions in the management of COVID-19.

## INTRODUCTION

Neutralizing autoantibodies (AABs) dysregulation is associated with clinical severity of severe coronavirus disease 2019 (COVID-19) disease. Bastard et al.^1,2^ characterized the presence of high titers of neutralizing AABs against interferons (IFNs), essential molecules for the immune response against viruses^3^ including the severe acute respiratory syndrome virus 2 (SARS-CoV-2), which increase the susceptibility to COVID-19-associated pneumonia and death. Further study showed diverse functional AABs were associated with severe COVID-19 infection, including those targeting cytokines (e.g., IL-1B and IL-6), chemokines (e.g., CCL11 and CXCL26), complement components (e.g., C5A and C9) and chemokine receptors (e.g., CCR2, CCRL2)^4^. These studies demonstrated the contribution of AABs to COVID-19 immunity.

We^5,6^ and other research groups^7–9^ reported that COVID-19 severity is also associated with the dysregulation of AABs associated with autoimmune diseases (*e.g.*, anti-phospholipid, anti-platelet glycoprotein, anti-nuclear AABs, and others). In particular, AABs against G protein-coupled receptors (GPCR) and renin-angiotensin system (RAS)-related molecules were associated with severe COVID-19 symptoms defined as requiring supplemental oxygen therapy^10^. These two groups of molecules are expressed by several human cells populations and modulate a myriad of intracellular signaling pathways and biological processes, such as cell trafficking, proliferation, survival, and differentiation, as well as neurotransmission and vasoconstriction^11–15^. Furthermore, several of these AABs act as functional, binding GPCRs and modulating intracellular pathways^16^ including those against the angiotensin receptor type 1 (AGT1R), which cause COVID-19-related symptoms^17,18^, such as skin and lung inflammation^19^. Notably, anti-AGT1R AABs have been implicated in various autoimmune conditions such as systemic sclerosis (SSc)^58^. There is evidence highlighting the pivotal role of endothelial dysfunction and injury in both SSc and COVID-19^59^. This endothelial cell activation and dysfunction represent a crucial and evolving step in the pathogenesis of these diseases^55^. However, while the mechanism of anti-AGTR1 AABs-induced pathology is better understood in SSc, in the context of COVID-19, the association of these AABs with disease development is not yet fully understood.

COVID-19 exhibits a diverse range of manifestations. Symptoms among individuals can vary widely, including fever, diarrhea, headache, depression, and amnesia^20^. The underlying causes of this variability remain elusive, particularly the potential role of AABs. This study aims to determine whether individuals with specific symptoms exhibit higher levels of certain AABs. Uncovering these associations could provide new insights into the pathophysiology of SARS-CoV-2 infections, which remains crucial. This is the case despite advancements in controlling the COVID-19 pandemic through prior infections, vaccinations^21^, and the increasing availability of treatments such as antivirals and immunomodulators^22^.

## METHODS

### Study cohort

We conducted a thorough investigation involving 244 unvaccinated adults residing in the United States. This cohort comprised 169 individuals diagnosed with COVID-19, confirmed through SARS-CoV-2 positive tests obtained via nasopharyngeal swab and polymerase chain reaction (PCR). Additionally, we included 75 randomly selected age, sex, and SARS-CoV-2 negative through PCR testing. COVID-19 patients were stratified according to the severity classification outlined by the World Health Organization (WHO). This categorization included mild cases (n=74), characterized by a fever duration of ≤ 1 day and a peak temperature of 37.8°C; moderate cases (n=63), exhibiting a fever duration of ≥ seven days and a peak temperature of ≥ 38.8°C; and severe cases (n=32), characterized by severe symptoms necessitating supplemental oxygen therapy. Every participant, including both healthy controls and patients, provided informed written consent in accordance with the principles set forth by the Declaration of Helsinki. The study received approval from the IntegReview institutional review board (Coronavirus Antibody Prevalence Study, CAPS-613) and adhered to the reporting guidelines outlined by Strengthening the Reporting of Observational Studies in Epidemiology (STROBE). Detailed demographic and clinical data are provided in the following sections (**Supplementary Table**).

### Measurements of anti-SARS-CoV-2 antibodies and AABs linked to autoimmune diseases

We detected human IgG AABs against 14 different GPCRs (AGTR1, AGTR2, MAS1, BDKRB1, ADRA1A, ADRB1, ADRB2, CHRM3, CHRM4, CHRM5, CXCR3, F2R, C5AR1), 2 molecules serving as entry for SARS-CoV-2 (ACE2, NRP1), and antibodies against the transmembrane receptor STAB1 from frozen serum using commercial ELISA kits (CellTrend, Germany) as previously described^10^. The assays were conducted according to the manufacturer’s instructions and as previously described. Briefly, duplicate samples of a 1:100 serum dilution were incubated at 4 °C for 2_Jh, and the AAB concentrations were calculated as arbitrary units (U) based on a standard curve of five standards ranging from 2.5 to 40 U/ml. The ELISA kits were validated following the Food and Drug Administration’s Guidance for Industry: Bioanalytical Method Validation.

### Enrichment of AAB targets

To perform functional enrichment analysis of the targets of AABs, we utilized the ClusterProfiler^23,24^ package in R^25^. This package enables the enrichment of gene sets that collectively participate in a common biological process (BP). For this purpose, we considered the 17 targets: ACE2, ADRA1B, ADRB1, ADRB2, AGTR1, AGTR2, BDKRB1, C5AR1, CHRM3, CHRM4, CHRM5, CHRNA1, CXCR3, F2R, MAS1, NRP1, and STAB1. Based on the results adjusted by false discovery rate (FDR), we only present the pathways that were deemed significant (adj p < 0.05). For the results visualization we used ggplot2^26^ R^25^ package and created with Biorender.com.

### Heatmap clustering and multi-study factor analyses

The levels of AABs and cohort features were visualized in a heatmap with hierarchical clustering (Euclidean distance) using the R^25^ packages ComplexHeatmap^27^ and Circlize^28^. Furthermore, we performed a multi-study factor analysis (MSFA) among AAB levels. The MSFA method integrates all data simultaneously, estimating parameters through maximum-likelihood analysis^29,30^. This approach allows for the identification of unobservable factors that may be specific (only in the control group and only in COVID-19 individuals and shared among the groups of healthy donors (Control) and COVID-19 cases.

### Classification of symptoms, relative effectors, and mixed canonical correlation analysis

We used the random forest model to rank the COVID-19 symptoms. Distinct groups were designated as 0 (non-COVID-19 disease, i.e., healthy donors) and 1 (COVID-19 disease, including mild, moderate, and severe cases). Furthermore, due to inherent imbalance in group sizes, the weight argument from the RandomForest^31^ R^25^ package was employed to correct this disparity and ensure balanced representation between groups^31^. After identifying the most relevant symptoms, subgroups were created to evaluate individuals with the presence of at least one or more of these key symptoms.

In addition, was performed the relative effects of AABs on four COVID-19 symptoms were assessed using a MANOVA analysis with bootstrap methodology, involving 1000 resamplings. Additionally, we computed confidence intervals (CI) for the relative effects using bootstrap techniques. The statistical analysis was performed using the R^25^ packages npmv^32^ and reshape2^33^. Finally, to demonstrate latent Gaussian correlation between binary variables (four symptoms individually) and continuous variables (AAB levels), we applied the mixed canonical correlation (CCA) with Kendall correlation and used the mixedCCA^34^ R^25^ package.

### Principal component analysis and factorial analysis

The Principal Component analysis (PCA) with spectral decomposition was conducted following previously outlined methods^5,35^. This approach allowed us to assess the discriminatory capacity of AABs in distinguishing symptom subgroups. To perform these calculations, we used specific R^25^ functions, namely get_eig and get_pca_var from the factoextra package^36^. The PCA itself was implemented using the prcomp function. Additionally, we employed latent factor analysis to identify sets of AABs (observed variables) that may indirectly interfere with the latent variables^37^. For this purpose, we used the factload function from the DandEFA^38^ R^25^ package.

### Median plots and multinomial logistic regression

We used median values to visually represent the distribution of significant levels of AABs in the presence and absence of symptoms in individuals with COVID-19. Statistical differences in AAB levels were assessed using the Kruskal-Wallis test, followed by post hoc analysis employing the Dunn test. Significance was established with a p-value < 0.05, and adjusted p-values FDR served as the significance cut-off. The creation of box plots was performed using the R packages rstatix^39^ and ggplot2^26^. In addition, the multinomial regression was applied to show AAB levels that decrease and increase through CI with odds ratio (OR), when compared with 0 group. For this we used the *multinom* function from nnet^40^ R^25^ package. The significance of the AABs was evaluated using a 95% exponential CI.

### Effects of anti-AGTR1 on the glycocalyx height & stiffness

To assess the height and stiffness of the endothelial glycocalyx (eGC), human umbilical vein endothelial cells (HUVECs) were cultured on coverslips until reaching confluence and then treated with either an anti-AGTR1 monoclonal antibody (anti-AGTR1 mAb, clone: 5.2a^19^) or the corresponding isotype control antibody (Purified Mouse IgG2a, clone: MG2a-53; BioLegend, San Diego, CA, USA) for 24 hours. In certain experiments, 1 µM Losartan (Losartan Carboxylic Acid, Cayman Chemical Company, Ann Arbor, MI, USA) was utilized to inhibit the AGTR1 function. Throughout the experimental procedure, cells were maintained in HEPES-buffered solution (Gibco, Waltham, MA, USA) supplemented with 1% fetal bovine serum (Bio&Sell, Feucht, Germany). The architecture of the eGC was evaluated utilizing Atomic Force Microscopy (AFM) nanoindentation technique (NanoWizard4, Bruker, Karlsruhe, Germany). This involved employing a triangular gold-coated cantilever with a spherical tip (diameter: 10 µm; Novascan, Ames, IA, USA) as the primary component of the AFM. The cantilever, with a spring constant of 10 pN/nm, was gently brought into contact with the cell surface with a maximal loading force of 0.5 nN to induce indentation. As the cantilever bent upon contact with the cell, the deflection of a laser beam focused on the surface of the gold-coated cantilever was monitored using a photodiode. Subsequently, force-distance curves were generated and analyzed to determine the height and stiffness of the glycocalyx. This analysis was facilitated by Protein Unfolding and Nano-Indentation Analysis Software (PUNIAS3D; Version 1.0; Release 2.3; Copyright 2009), followed by statistical evaluation of the obtained values. Statistical analysis was performed using R^25^ programming. Shapiro Wilk test was applied to test for normality of the data distribution. To compare the means of the non-parametric groups, Kruskal Wallis test was used, followed by Dunn’s post-hoc test for multiple comparisons. Data are presented as mean ± standard error of the mean (SEM) and FDR < 0.05 was considered statistically significant.

## RESULTS

### Serum AAB signatures associated with COVID-19 symptoms

Initially, hierarchical clustering association between serum levels of AABs targeting 17 receptors, selected based on their functionally and hypothesized association with COVID-19 symptoms (**Figure 1a**). I.e., these receptors include GPCRs not previously associated with the RAS (CHRM3, CHRM4, CHRM5, CHRNA1, CXCR3, C5AR1, F2R, NRP1, STAB1), as well as molecules belonging to or influencing the RAS (herein referred to as RAS-related molecules). The latter category comprises non-GPCRs (ACE2 and MAS1) as well as GPCRs (ADRB1, ADRB2, AGTR1, AGTR2, ADRA1A, and BDKRB1). These molecules are expressed in various body tissues, including those of the nervous, circulatory, and immune systems^41^. They are associated with biological processes relevant to COVID-19 pathophysiological mechanisms, including vasculopathy, cognitive dysfunction, and hyper inflammation. (**Figures 1b**).

**Figure 1:**
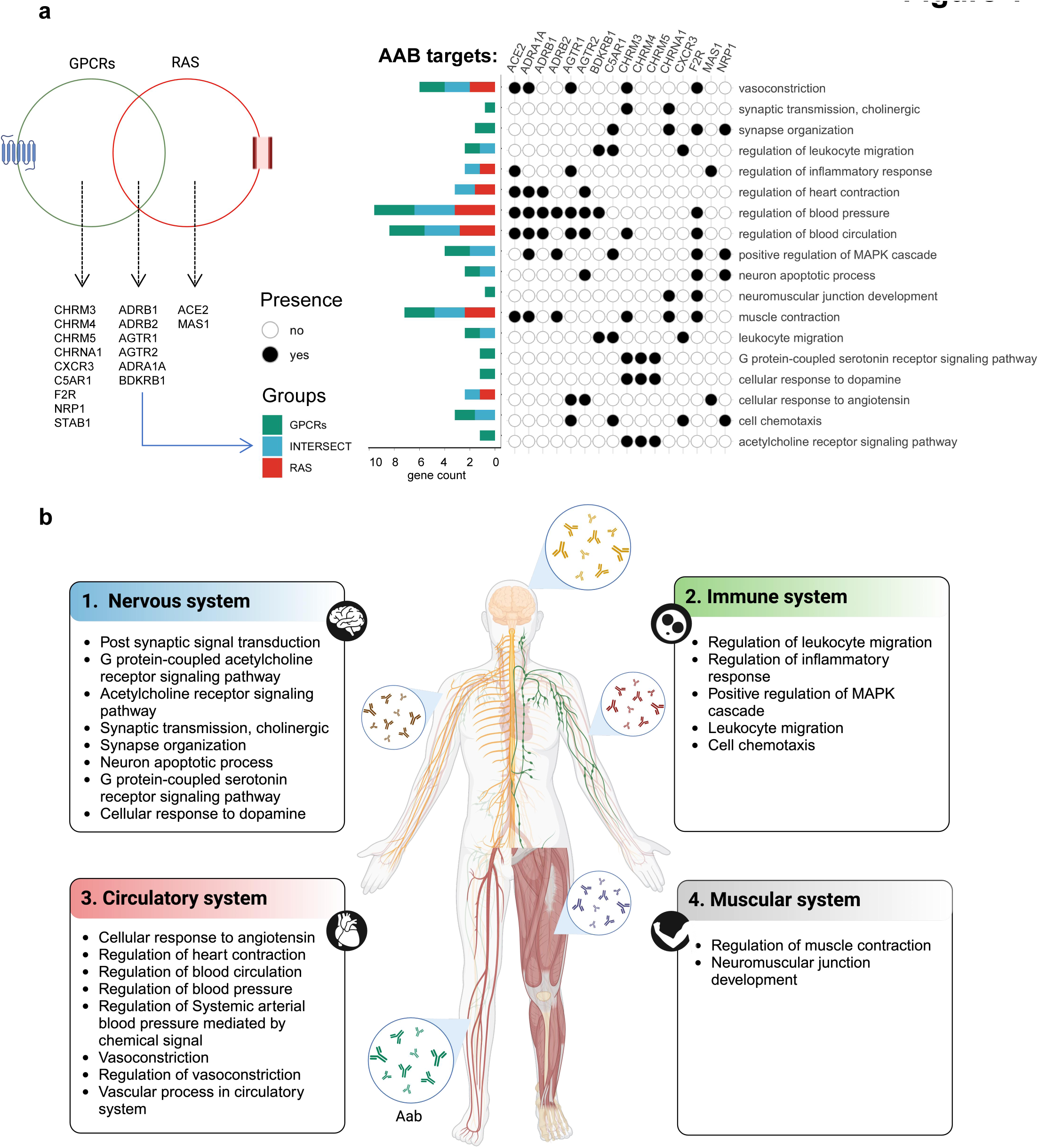
Biological processes linked to antibody targets. **a)** The Venn diagram shows the AAB targets belonging to either the GPCR or RAS group. Only gene sets present in significant pathways according to the FDR are shown. Additionally, the graphic on the left exhibits the enriched biological processes (BPs) associated with these AAB targets, and **b)** the different systems to which these BPs are linked.

The hierarchical clustering of the AABs revealed a consistent trend: as the severity of COVID-19 progressed from mild to moderate to severe disease, there was a corresponding increase in the levels of these antibodies, along with their association with the disease symptoms. The number of patients with each symptom is described in **Supplementary Figure 1**. For example, symptoms like muscle ache and fever displayed a higher propensity to cluster in line with moderate and severe COVID-19 patients than other symptoms, such as diarrhea and dysgeusia (**Figure 2a**). Here, the control group is formed by healthy individuals or SARS-CoV-2 negative controls presenting at least one symptom of gastro-intestinal or respiratory disease. Of note, given that we recently characterized our COVID-19 cohort^6,10,42,43^, finding them to have comparable average age, sex distribution, and sample collection dates, we excluded these variables as potential confounders in our analyses.

**Figure 2:**
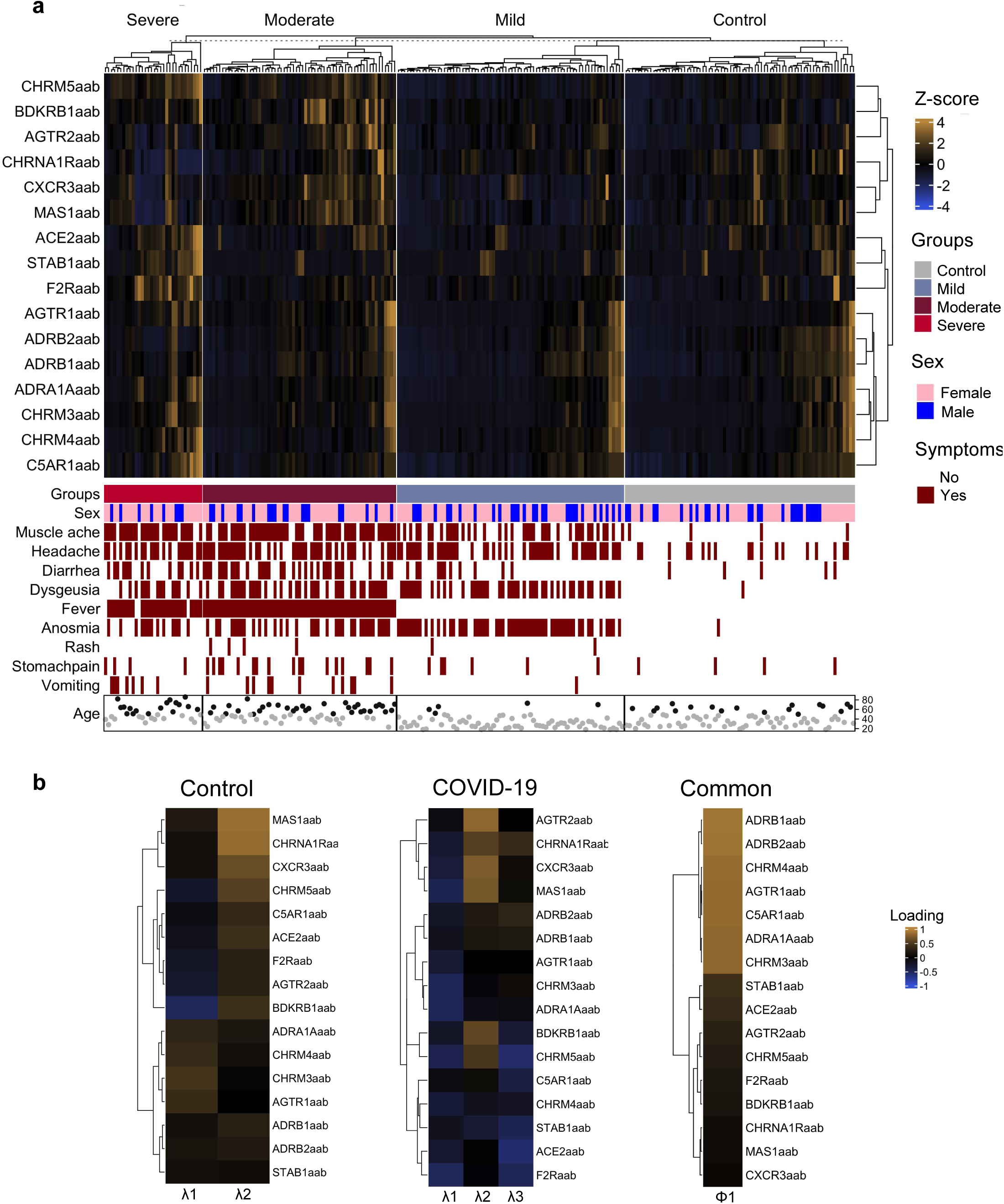
AAB levels according to COVID-19 severity and symptoms. **a**) A hierarchical clustering heatmap displays AAB levels (after z-score transformation), indicated by the scale bar. The presence or absence of symptoms, sex, and age range categories (<50 and ≥50, represented by gray and black circles) are shown below the heatmap. **b)** Multistudy factor analysis of AABs. Heatmaps shows the result of multistudy factor analysis (MSFA), indicating estimated factor loadings for common and specific latent factors between controls and COVID-19 patients. The color scale bar ranging from orange (−1 to 1) to blue corresponds to negative and positive factor loadings. Loadings close to −1 or 1 indicate aab that strongly influence factors in opposite directions.

Furthermore, MSFA indicated the presence of specific latent factors when comparing healthy controls to COVID-19 patients (**Figure 2b**), supporting our previous hypothesis of AABs playing a role in both health and disease. In this context, the existence of common (shared) latent factors suggests physiological functions that are regulated by AABs but are not affected by the disease state.

### Identifying key symptoms and AABs in COVID-19 using random forest analysis and relative effect

To identify the most relevant symptoms within our cohort (**Figure 3a**) related to the COVID-19 phenotype and to further explore the correlation between the concentration of AABs and these symptoms, we employed random forest analysis, which is a random forest approach. This method is capable of identifying the most significant predictors of a given phenotype^44^. Our analysis indicated that anosmia, muscle ache, fever, and dysgeusia were the most relevant symptoms defining our COVID-19 cohort (**Figure 3b**). The random forest model achieved an area under the curve (AUC) above 70% for both specificity and sensitivity for each symptom (**Figure 3c**).

**Figure 3:**
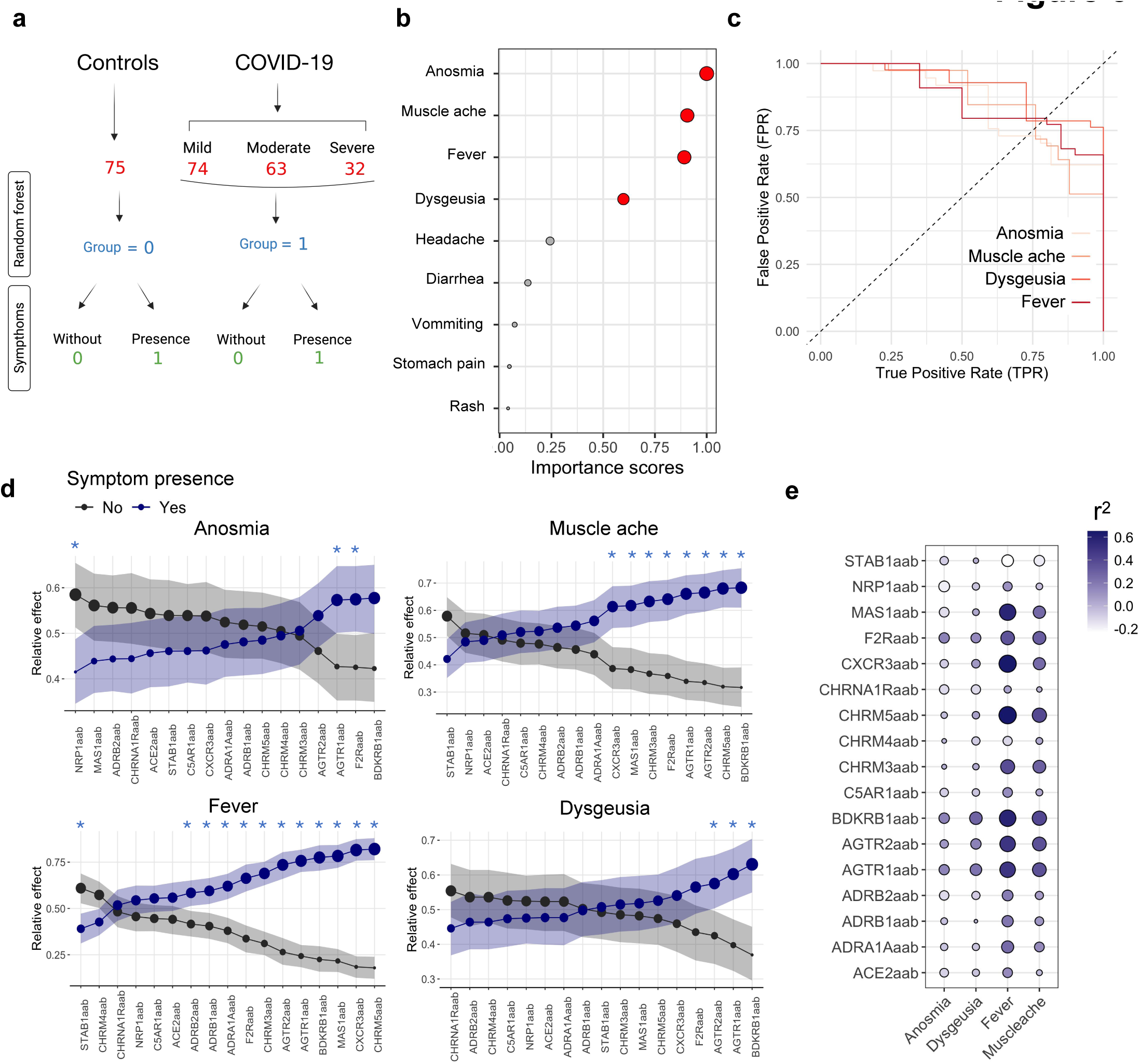
Relative effect of aabs on COVID-19 symptoms. **a)** A flowchart of COVID-19 severity and healthy groups as labeled for random forest analysis. The control group consists of healthy individuals or SARS-CoV-2 negative controls presenting at least one symptom of gastrointestinal or respiratory disease. They were classified as individuals without (0) or with the presence of symptoms (1). **b)** A dot plot portrays the importance score from random forest analysis of COVID-19 symptoms. Red dots represent symptoms with an importance score above 0.5 (50%). The circle size increases according to the importance score. **c)** ROC curves show the False Positive Rate (FPR) and True Positive Rate (TPR) for symptoms with the highest importance scores (anosmia, muscle ache, fever, and dysgeusia). **d**) The relative effects were calculated using MANOVA test. The circle size indicates the probabilistic measure (relative effect size). Confidence intervals are shown by shadows. Black and blue lines/dots represent individuals with and without symptoms, respectively. Furthermore the significance interval is identified by blue asterisks. **e)** The bubble heatmap shows the canonical correlation results. The size and color spectrum of the bubbles represent the r2 value between symptoms and AAB levels.

Next, we investigated which AABs could be associated with the development of the four most relevant symptoms that predict the phenotype of our COVID-19 cohort. This determination was based on our analysis of the correlation strength between these AABs and the symptoms, alongside other relevant factors considered in our study methodology. We conducted a relative effector analysis using the bootstrap and MANOVA test, which serves as a probabilistic measure to assess the likelihood of AABs influencing COVID-19 symptoms. This approach revealed distinct patterns of AABs behavior and their associations with each symptom (**Figure 3d**). Specifically, AABs targeting F2R, AGTR1, and NRP1 showed the strongest association with anosmia, while those against BDKRB1, CHRM5, AGTR2, and AGTR1 were most closely linked to muscle ache. Additionally, anti-CHRM5, anti-CXCR3, anti-MAS1, and CHRM5 displayed the highest correlation with fever, and anti-BDKRB1, anti-AGTR1, anti-AGTR2, and anti-F2R were most strongly associated with dysgeusia. In agreement, CCA indicated AABs targeting AGTR1, AGTR2, BDKRB1, CHRM3, CHRM5, CXCR3, F2R, and MAS1 with high positive correlation with at least one symptom (**Figure 3e**).

### Exploring AAB profiles as predictors of COVID-19 symptom severity

To further investigate the correlations between AABs and the four most relevant symptoms predicting the phenotype of our COVID-19 cohort, we explored the stratification capacity of AABs levels based on the accumulation of COVID-19 symptoms. To achieve this, we conducted PCA with spectral decomposition using the AABs levels of controls and COVID-19 patients. We categorized these individuals into five groups based on the number of symptoms present and their importance score (>50 or <50) (**Figure 3b and 3c**). Therefore, healthy controls, other controls without COVID-19 but with symptoms of mild respiratory illness, and a unique severe COVID-19 patient who, at the time of sample collection, neither exhibited the most relevant symptoms defining our COVID-19 cohort (anosmia, muscle ache, fever, and dysgeusia) nor the other symptoms considered in our analysis due to extreme illness and, were categorized as *group 0*. Since we did not observe striking differences in the hierarchical clustering pattern of AABs when excluding the other controls without COVID-19 but with symptoms of mild respiratory illness and this unique severe COVID-19 group (**Supplementary Figure 2**), we assumed that they did not impact the global pattern of the results obtained. Finally, we defined COVID-19 patients exhibiting one, two, three, or four symptoms as *groups 1, 2, 3*, or *4* (**Supplementary Figure 3**).

This approach revealed that AABs progressively stratify COVID-19 patients (**Figure 4a**). In essence, as the number of symptoms (anosmia, dysgeusia, muscle ache, and fever) increases, there is an observable trend of higher levels of these AABs in patients. Specifically, individuals presenting all four symptoms show a more distinct separation from healthy controls compared to those with three, two, or one of these symptoms. Notably, the AABs identified through the relative effect analysis were found to be part of two major contribution clusters that appeared to play a significant role in stratifying COVID-19 symptoms (**Figure 4b**). The first cluster, comprising CXCR3-AABs, CHRM5-AABs, BDKRB1-AABs, and MAS1-AABs, was shown to contribute most significantly to the principal component (PC) 2. The second cluster, which had a greater impact on PC1, included AGTR1-AABs, ADRA1-AABs, CHRM3-AABs, C5AR1-AABs, ADRB1-AABs, ADRB2-AABs, and CHRM4-AABs. A third cluster of AABs, with less contribution to both PC1 and PC2, consisted of AGTR2-AABs, CHRNA1, F2R-AABs, STAB1-AABs, NRP1-AABs, and ACE2-AABs.

**Figure 4:**
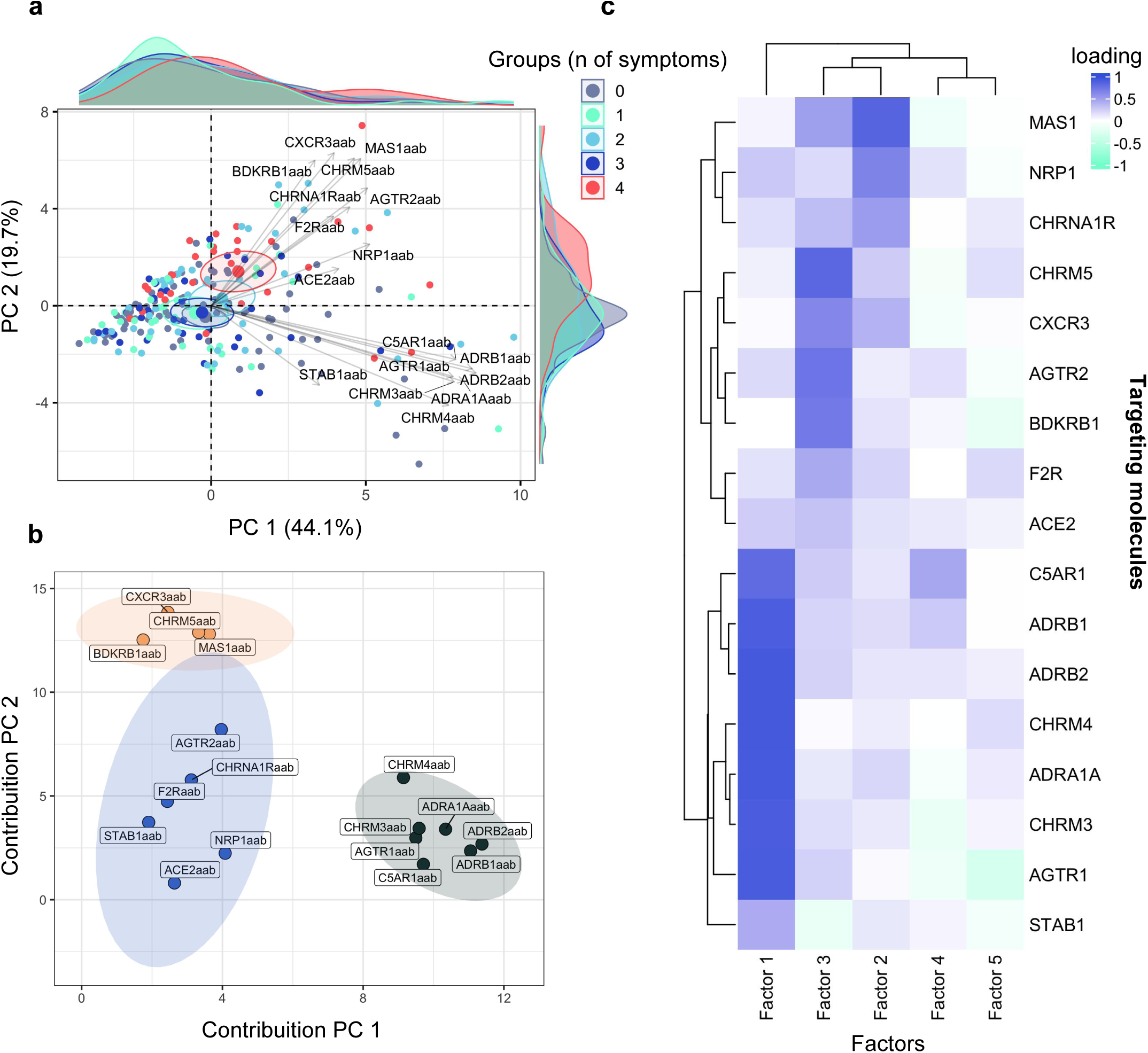
AABs stratify COVID-19 according to symptoms. **a)** The PCA graphic illustrates the stratification of COVID-19 groups based on the number of symptoms (0 = no symptoms; 1 = one symptom; 2 = two symptoms; 3 = three symptoms; 4 = four symptoms). Variables with positive correlation are plotted on the same side, while negatively correlated variables are plotted on opposite sides. Only AABs that highly contribute to stratifying moderate and severe COVID-19 patients from mild patients and healthy controls are displayed. Small circles represent concentration ellipses around the mean points of each group. Histograms alongside the PCA plot represent the density of the sample (individual) distribution. **b)** The scatter plot shows the contribution of variables (AABs clusters 1, 2, and 3 are represented by the colors black, blue, and orange, respectively) to dimensions 1 and 2. These variables indicate the contribution of AABs to the group stratification. **c)** The heatmap displays factorial load values between AABs, indicating the strength of the relationship (ranging between 1 and -1) between the variable and the factor.

Additionally, we observed a similar clustering tendency in the AABs profile, where those targetingCHRM5, CXCR3, AGTR2, BDKRB1, and F2R had the highest contribution score for factor 3, while AABs against C5AR1, ADRB1, ADRB2, CHRM4, ADRA1, CHRM3, and AGTR1 showed the highest contribution score for factor 1 in our EFA (**Figure 4c**). The goal of EFA is to uncover underlying structures or factors that explain the correlations among a set of observed variables. These factors are latent constructs that cannot be directly measured but are inferred from the patterns of correlations among the observed variables^45^. This result reinforces the possibility that AABs are contributing synergistically to COVID-19 symptoms, possibly through their dysregulation of the biological processes in which they are involved.

### Association of dysregulated AABs with COVID-19 symptom accumulation

The stratification results described above suggest that levels of AABs dysregulate with the accumulation of COVID-19 symptoms. To address this, we conducted a multiple comparison analysis between patients without symptoms and those presenting one, two, three, or all four symptoms (anosmia, dysgeusia, muscle ache, and fever). This approach revealed significant alterations in several AABs with COVID-19 symptoms accumulation, namely, AABs targeting ACE2, AGTR1, AGTR2, BDKRB1, CHRM3, CHRM5, CXCR3, F2R, and MAS1 (**Figure 5a**).

**Figure 5:**
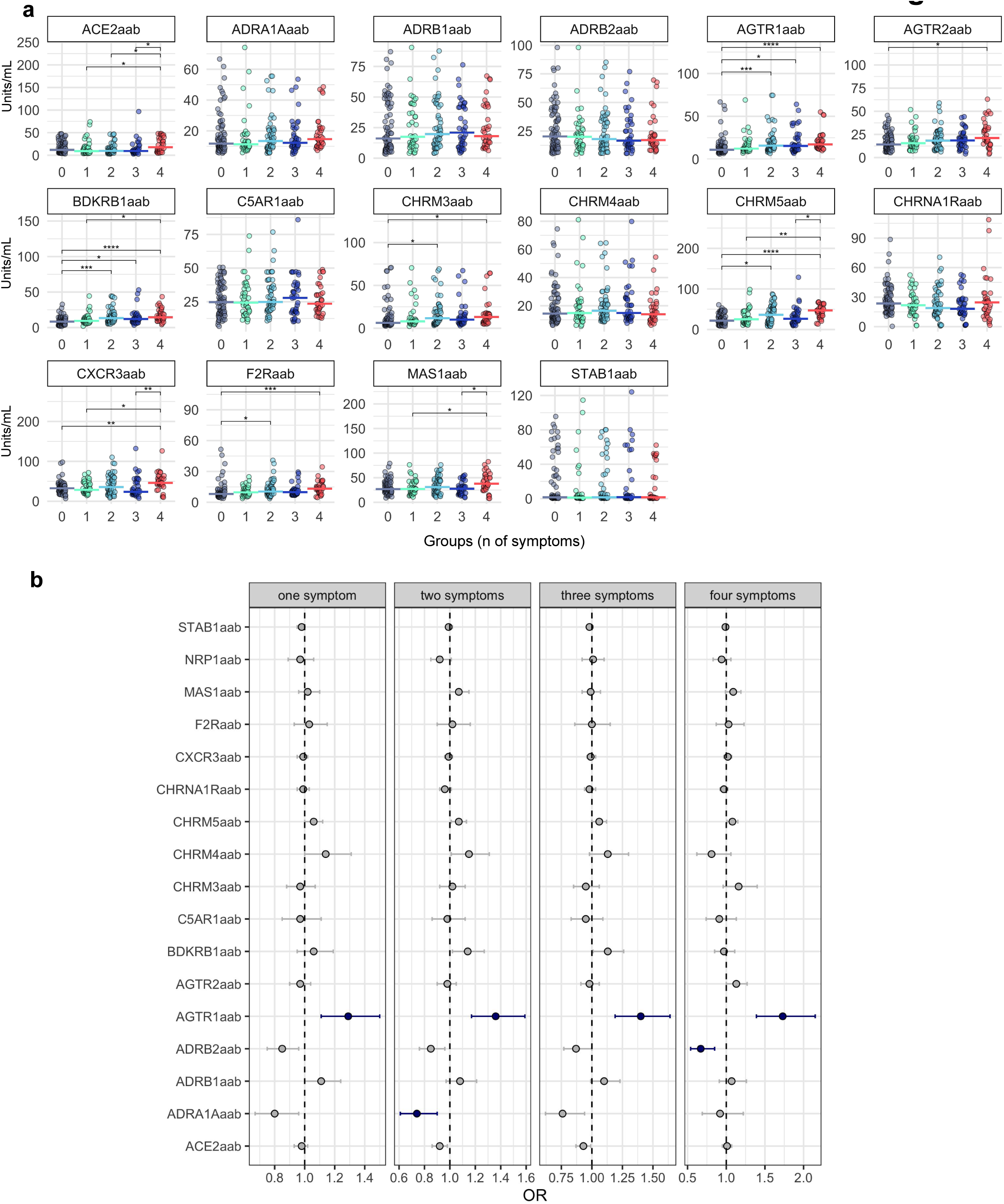
Functional effects of anti-AGTR1 on the glycocalyx height & stiffness. **a)** Dose response curve of anti-AGTR1 antibodies or isotype control on heights and stiffness of glycocalyx. **b)** Decreased endothelial glycocalyx (eGC) height and elevated stiffness after treatment with anti-AGTR1 antibodies is reversed by Losartan.

Given that multiple comparisons, such as Dunn’s test following Kruskal-Wallis, increase the risk of Type I error (false positives), we further conducted a multinomial logistic regression analysis. This analysis inherently adjusts for multiple comparisons, potentially providing a more conservative estimate of significance. This approach was performed to evaluate the association between AAB levels and the accumulation of COVID-19 symptoms more rigorously. It revealed that the anti-AGTR1 antibody was the only one strongly associated with significant OR and FDR adjusted p-values with the development of one, two, three, or all four of the assessed symptoms (**Figure 5b**).

### Functional effects of anti-AGTR1 on the glycocalyx height & stiffness

Inflammation-induced degradation of the eGC, a critical component in preserving endothelial function, has been implicated in the pathogenesis of COVID-19-related endothelial dysfunction^46–48^. Utilizing an anti-AGTR1 monoclonal antibody (mAb) across various concentrations (10, 50, and 100 µg/mL), we observed concentration-dependent reductions in eGC height and increases in stiffness compared to isotype controls (**Figure 6a**). Notably, even at the lowest concentration (10 µg/mL), the anti-AGTR1 mAb significantly diminished eGC height by approximately 25% and heightened stiffness by over 50% (p<0.0001). Incremental concentrations exacerbated these effects, with a peak reduction in eGC height and increased stiffness at 50 µg/mL.

**Figure 6:**
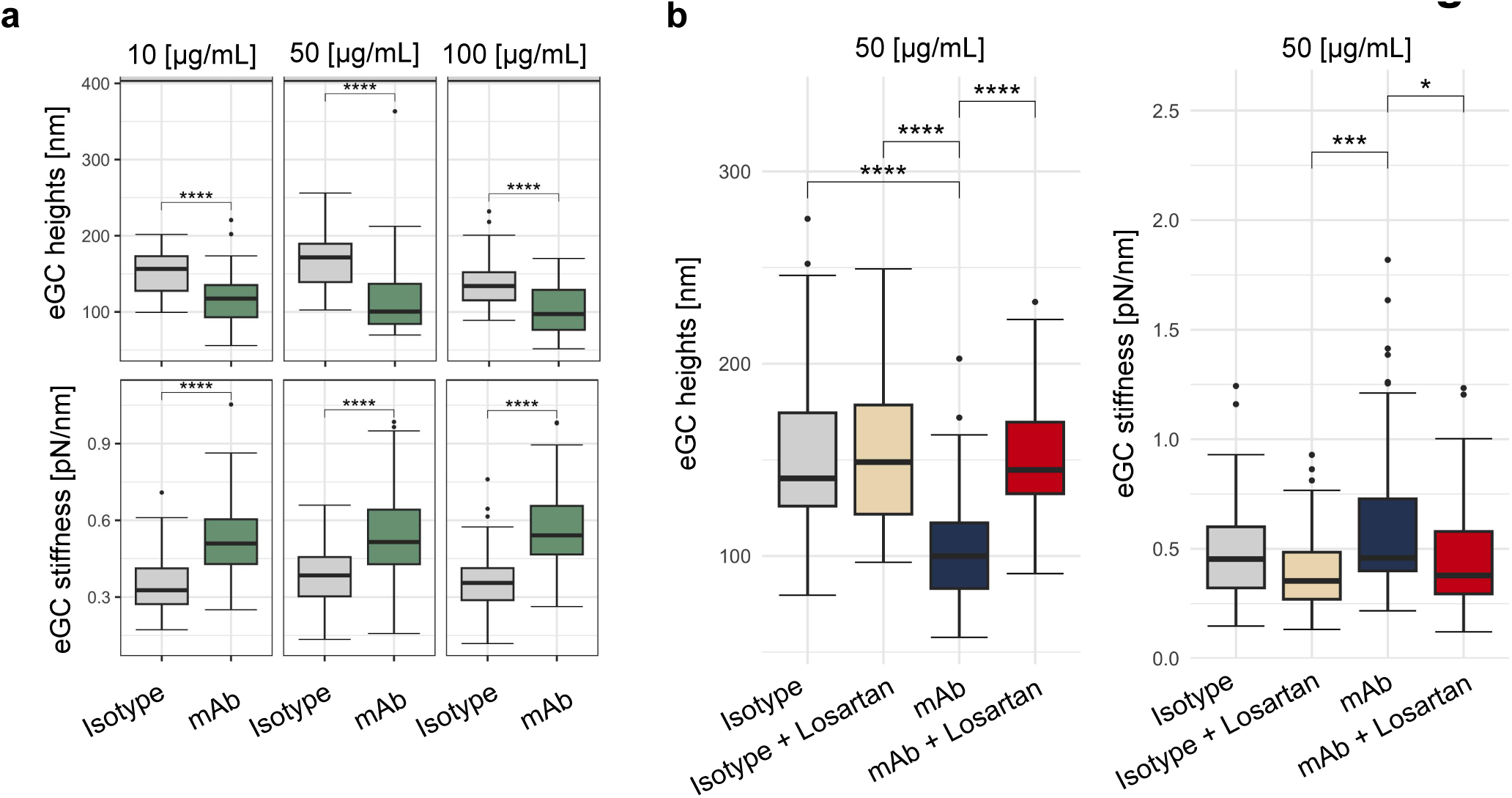
AAB levels dysregulate with the accumulation of COVID-19 symptoms. **a)** Violin plots display AAB levels for each group in the x-axis (0 = no symptoms; 1 = one symptom; 2 = two symptoms; 3 = three symptoms; 4 = four symptoms). Asterisks indicate the Kruskal-Wallis test with post-hoc Dunn test significance levels (* = p < 0.01; ** = p < 0.001; *** = p < 0.0001; **** = p < 0.00001). Adjusted *p*-values (FDR) are also shown. **b**) Forest plots depict odds ratios (ORs) and their corresponding 95% confidence intervals (whiskers) for various AABs across COVID-19 symptoms. Blue dots and lines indicate significantly increased or decreased AAB levels compared to those in healthy controls. This significance is based on FDR and CI. The dashed line represents the intercept.

To assess the specificity of anti-AGTR1 mAb actions and explore potential protective strategies, we employed Losartan, an AGTR1 antagonist, in conjunction with the mAb. Treatment with anti-AGTR1 mAb (50 µg/mL) plus Losartan restored eGC height by nearly 45% and reduced stiffness by about 18% relative to anti-AGTR1 mAb treatment alone, underscoring the protective effect of Losartan against eGC degradation (**Figure 6b**). Controls with Losartan alone showed no significant changes, confirming the specificity of the anti-AGTR1 mAb effects.

## DISCUSSION

This manuscript presents a comprehensive analysis of AABs targeting GPCRs and RAS-related molecules in relation to COVID-19 symptoms. The hierarchical clustering analysis revealed an increasing trend in AABs levels corresponding to the severity of COVID-19 and associated symptoms. The machine learning approach identified anosmia, muscle ache, fever, and dysgeusia as the most relevant symptoms defining the COVID-19 cohort, underscoring the importance of these symptoms in characterizing COVID-19. The relative effect and CCA analysis further elucidated the association between specific AABs and COVID-19 symptoms. For instance, AABs targeting NRP1, F2R, and AGTR1 were strongly associated with anosmia, while those against BDKRB1, CHRM5, AGTR2, and AGTR1 were linked to muscle ache. The stratification analysis based on the accumulation of COVID-19 symptoms demonstrated that AABs progressively stratify COVID-19 patients, with those presenting all four relevant symptoms showing a clearer separation from healthy controls. The analysis of dysregulation of AABs levels with the accumulation of COVID-19 symptoms further strengthens the association between these AABs and COVID-19 pathophysiology. Although further studies are warranted to validate these findings and explore the potential therapeutic implications of targeting these AABs in COVID-19 management, our data provide valuable insights into the role of AABs targeting GPCRs and RAS-related molecules in COVID-19 pathophysiology and symptomatology. This study focused on COVID-19 symptomatology adds a new dimension to understanding COVID-19 pathophysiology by providing a comprehensive analysis of AABs targeting GPCRs and RAS-related molecules.

Notably, AABs targeting GPCRs is an evolving history in autoimmunity^49^ and they functionally have been well characterized^49^. For instance, anti-AGT1R AABs, which showed the strongest association with the accumulation of COVID-19 symptoms, have been shown to trigger *in vitro* and *in vivo* effects also developed by COVID-19 patients^50–53^, such as lung hyperinflammation, infiltration of immune cells, and endothelial damage^19,54,55^. Apart from its role in the renin-angiotensin system, angiotensin II also exhibits pro-inflammatory effects by stimulating ADAM metallopeptidase domain 17 (ADAM17), leading to the production of inflammatory cytokines such as INF-γ, IL-8 and interleukin-6^54,56,57^. A recent study has demonstrated that anti-AT1R antibodies can act in an agonistic and synergistic manner with angiotensin II^19^. Hence, these antibodies could potentially enhance the effects of angiotensin II, contributing to the development of COVID-19 symptoms. Nevertheless, since compelling emerging data suggest that anti-AGTR1 AABs may play a role in the pathophysiology of COVID-19^10,56,60,61^, we hypothesize that these AABs might contribute to the dysregulation of the RAS, promote hyperinflammation, and be implicated in the endothelial dysfunction presented by COVID-19 patients as they are involved in the etiopathogenesis of SSc. However, further research is needed to validate these findings and understand the underlying mechanisms by which anti-AGTR1 AABs contribute to COVID-19 pathophysiology and the precise mechanisms and clinical implications of anti-AT1R AABs in COVID-19.

Moreover, decreased eGC height and elevated stiffness after treatment with anti-AGTR1 antibodies being reversed by Losartan, indicates a possible new specific pathological effect of antiAGTR1 AABs. This result is in agreement with emerging evidence suggesting that the degradation of the eGC^62^, a key regulator of vascular homeostasis^63^, plays a critical role in the constellation of COVID-19 symptoms^48,60^. The eGC’s impairment, as indicated by our findings, could contribute to systemic manifestations such as anosmia and dysgeusia^64^. These sensory deficits may arise from compromised microvascular integrity within the olfactory and gustatory systems, leading to disrupted cellular function in these regions. Additionally, the observed increase in eGC stiffness and reduced height may impede muscle perfusion, potentially elucidating the myalgia experienced by many COVID-19 patients^65^. Fever, a hallmark of the body’s inflammatory response to infection^66^, may also be potentiated by eGC damage^67^. The resultant endothelial dysfunction could amplify cytokine production and release, precipitating the febrile response. Together, these associations underscore the need for further investigation into the impact of eGC degradation on vascular health and its implications for the multisystemic symptoms encountered in COVID-19, potentially offering novel insights into targeted therapeutic interventions.

In conclusion, the comprehensive analysis presented in this work provides crucial insights into the nuanced interactions between AABs and specific COVID-19 symptoms, shedding light on the differential associations observed across varying symptomatology. For instance, the hierarchical clustering analysis showed an increasing trend in AABs levels corresponding to COVID-19 severity and its associated symptoms, with specific AAB strongly linked to symptoms such as anosmia and muscle ache. The study also highlighted the progressive stratification of COVID-19 patients based on autoantibody levels and the dysregulation of these levels with the accumulation of symptoms. Importantly, this study is the first to investigate the association between these AABs and specific COVID-19 symptoms, adding a new dimension to our understanding of COVID-19 pathophysiology. Further research is needed to validate these findings and explore the therapeutic implications of targeting these AABs in COVID-19 management.

## Data availability

All data used in this study are provided in the Supplementary Data.

## Author Contributions

DLMF, RA, and OCM wrote the manuscript; DLMF, ISF, OCM, RA, MJ, SLS, BF, CV, XW, LFS, RJSD, HDO, GCM, RC, GCB, PPF, YRAY, HN, RFC, MHH, GM, RC, FYNV, YLGC, AHCM, JNU, ALN, ASA, PBM, TSS, JCSS and LOL provided scientific insights; DLMF, OCM, GCB, PPF, and AHCM performed data and bioinformatics analyses; HDO, GM, RC, OCM, LFS, YRAY, GCM, RC, RA, KKV, and, AHCM revised and edited the manuscript; DLMF, AHCM, HH, AV, HA, IZ, AZR, GR, RA, KKV, YS, KSF, and OCM conceived the project and designed the study; JIS, AZR, and IZ diagnosed, recruited or followed-up the patients; KSF, HH, AZR, GH, YO, JZ, JIS, IZ, EV, YBL and YS coordinated the serum collection and databank or performed the experiments; IZ, AZR, AV, YS, AHCM, GR, RA, and OCM supervised the project.

## Data Availability

All data produced in the present work are contained in the manuscript

## Acknowledgments

We acknowledge the patients for participating in this study. We would like to recognize the contributions of Lev Rochel Bikur Cholim of Lakewood (led by Rabbi Yehuda Kasirer and Mrs. Leeba Prager) and the hundreds of volunteers who collected samples for this research through the MITZVA Cohort. We thank the São Paulo Research Foundation (FAPESP grants 2018/18886-9, 2020/01688-0, and 2020/07069-0 to OCM, 2020/16246-2 and 2023/13356-0 to DLMF, 2020/09146-1 to PPF, 2020/07972-1 to GCB, 2023/12268-0 to ASA, 2023/06086-6 to PMB, and 2019/27139-5 to JCSS) for financial support. We acknowledge the National Council for Scientific and Technological Development (CNPq) Brazil (grants: 309482/2022-4 to OCM and 102430/2022-5 to LFS) and the the Coordination of Superior Level Staff Improvement (CAPES) (grants: 88887.801068/2023-00 to ALN, CAPES/PROEX grant 88887.917898/2023-00 to JNU and 88887.699840/2022-00 to FYNV). The contributions by G.M. and R.C. were made possible by funding from the German Federal Ministry for Education and Research (BMBF) and German Research Foundation (DFG; projects #394046635, subproject A03, as part of CRC 1365, and EXPAND-PD; CA2816/1-1) through the Berlin Institute of Health (BIH)-Center for Regenerative Therapies (BCRT) and the Berlin-Brandenburg School for Regenerative Therapies (BSRT, GSC203), respectively, and in part by the European Union’s Horizon 2020 Research and Innovation Program under grant agreements No 733006 (PACE) and 779293 (HIPGEN). We also acknowledge the German Research Foundation (DFG: Research Training Group ’Autoimmune Pre-Disease’ RTG 2633 and Excellence Cluster ’Precision Medicine in Inflammation’ EXC 2167) and the German Ministry of Education and Research (BMBF, Mesinflame consortium no. 01EC1901D).

---

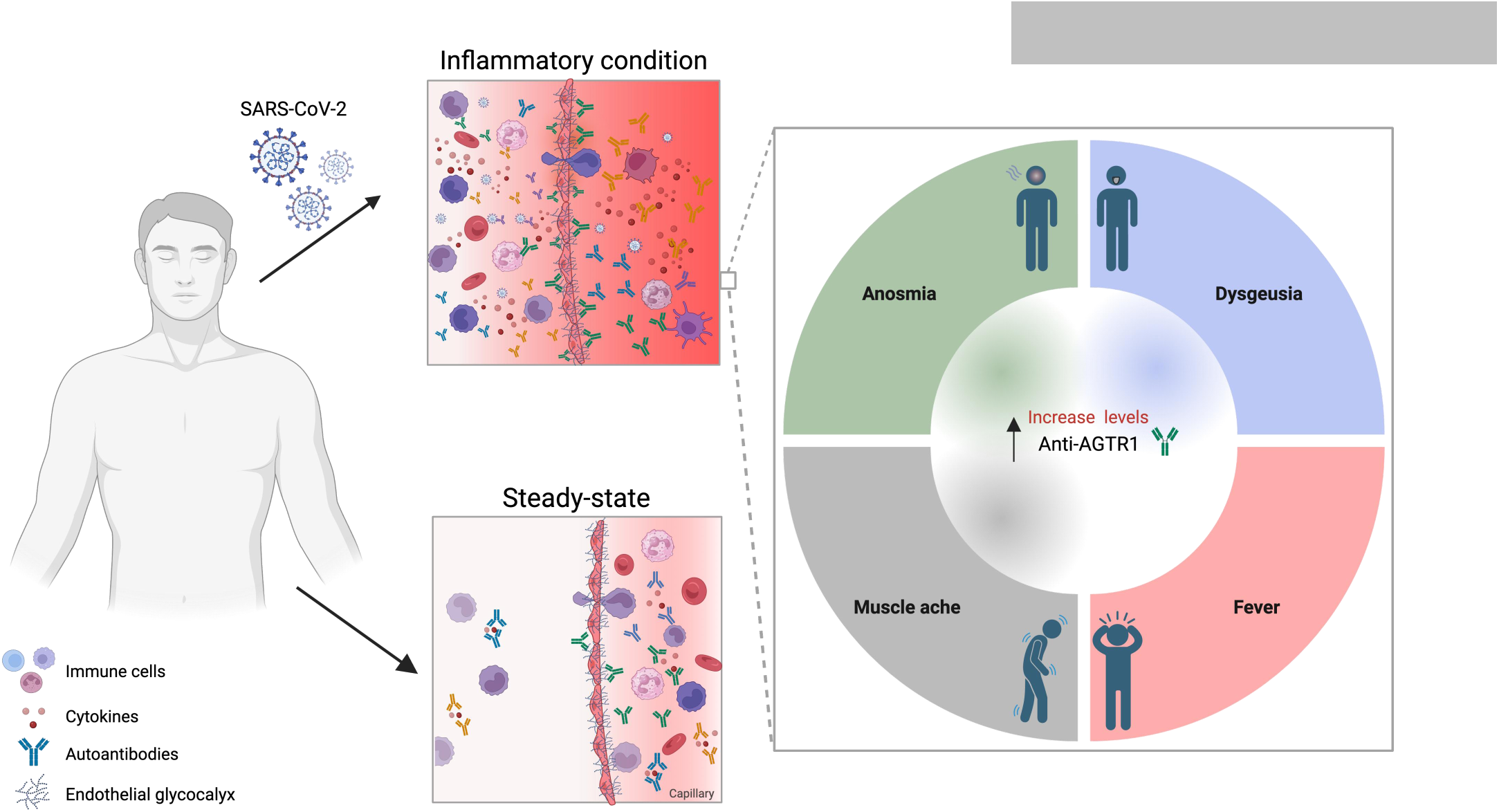
Graphical abstract

**Supplementary Figure 1:**
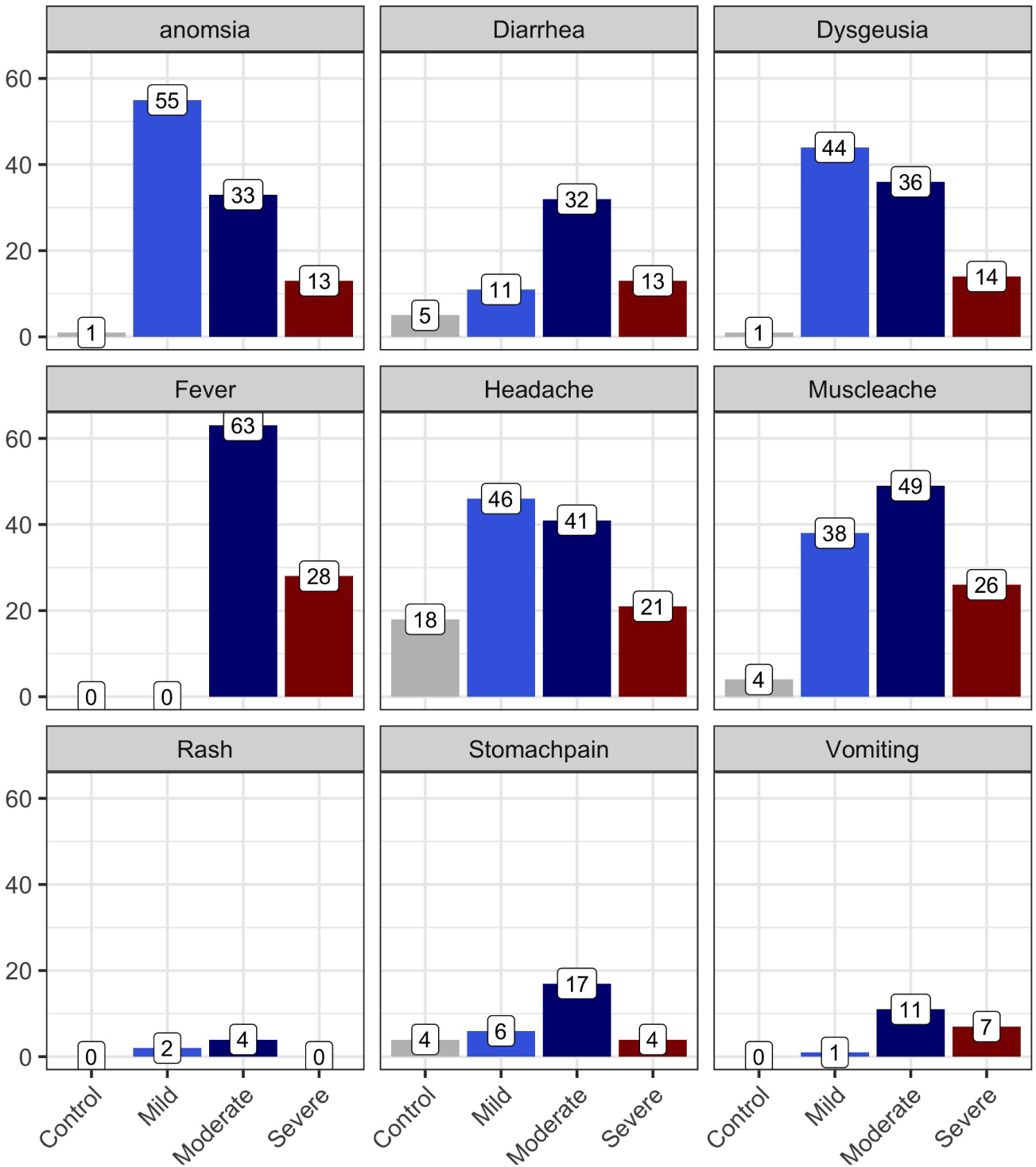
Number of patients with symptoms in COVID-19 severity groups.

**Supplementary Figure 2:**
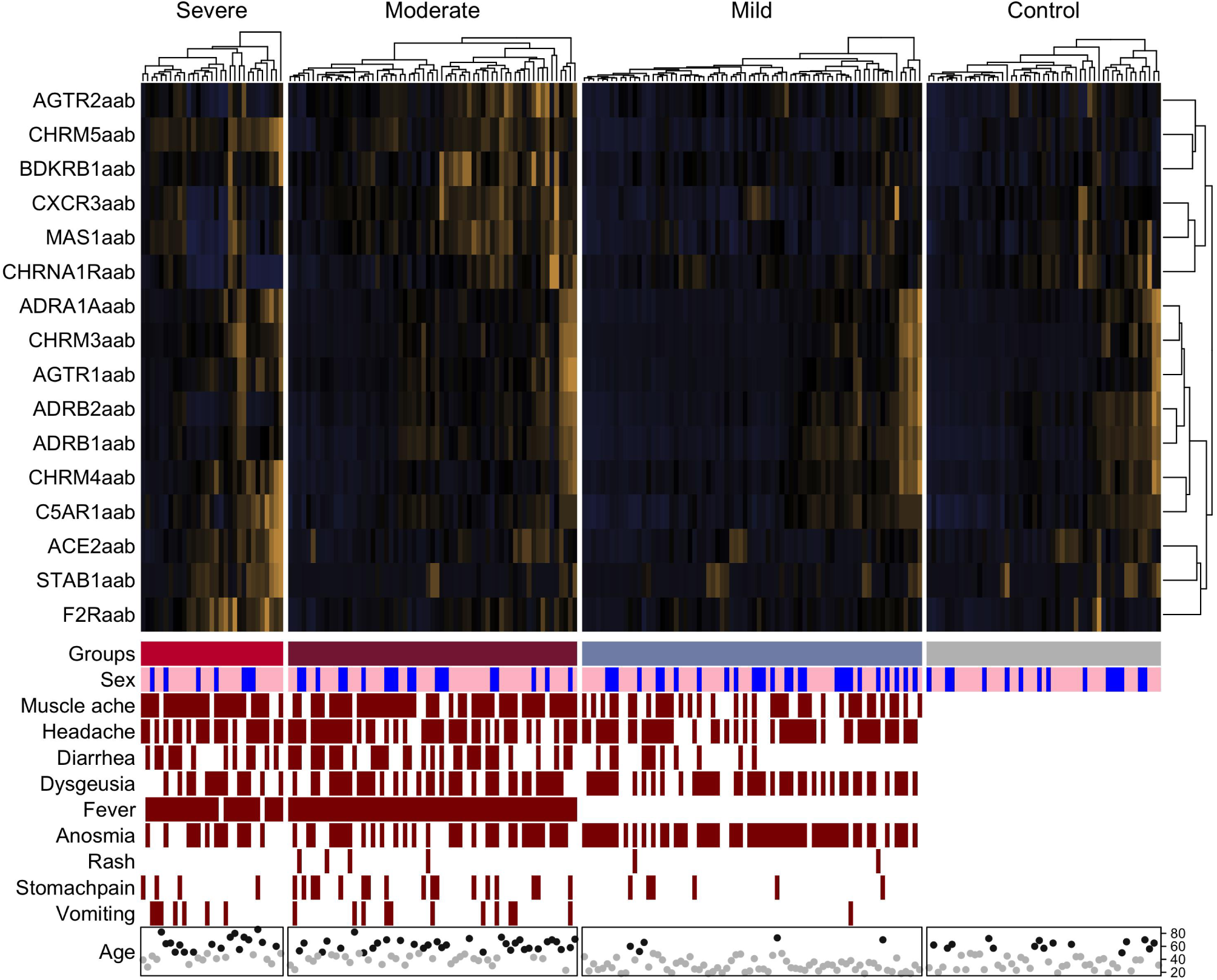
Heatmap showing AAB levels (after z-score transformation), indicated by the scale bar. The presence or absence of symptoms, sex, and age range categories (<50 and ≥50, represented by gray and black circles), are displayed below the heatmap. This heatmap excludes healthy individuals with symptoms and the patient in the severe group without any symptoms.

**Supplementary Figure 3:**
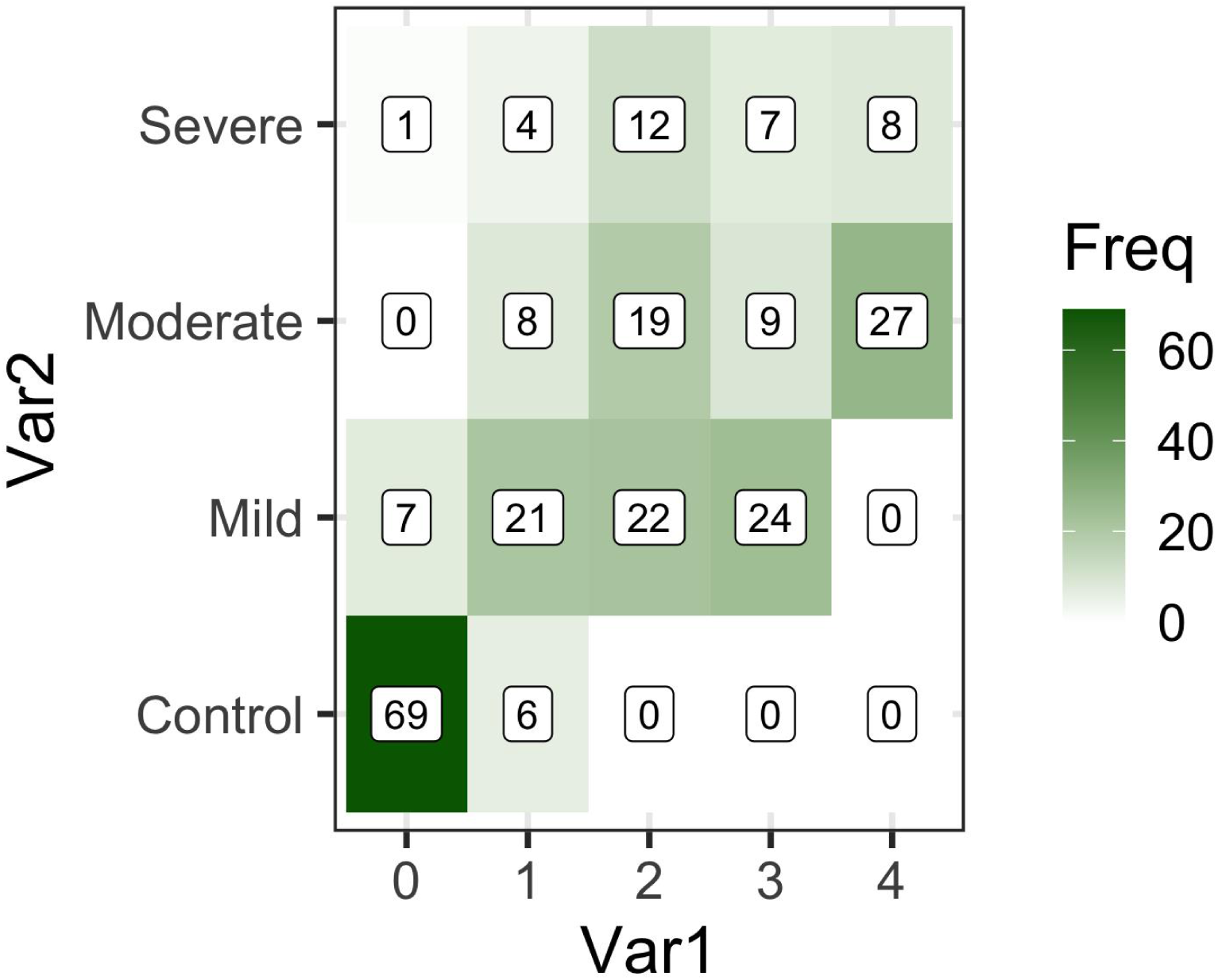
The heatmap categorizes individuals into groups 0, 1, 2, 3, and 4 based on the PCA analysis (Figure 5a and Figure 6), illustrating the distribution of individuals in each group.

